# Metapopulation modeling of COVID-19 advancing into the countryside: an analysis of mitigation strategies for Brazil

**DOI:** 10.1101/2020.05.06.20093492

**Authors:** Guilherme S. Costa, Wesley Cota, Silvio C. Ferreira

## Abstract

Since the first case of COVID-19 was confirmed in Brazil on 19 February 2020, this epidemic has spread throughout all states and at least 2142 of 5570 municipalities up to 30 April 2020. In order to understand this spreading, we investigate a stochastic epidemic model using a metapopulation approach. Simulations are supplied with real data for mobility, demography, and confirmed cases of COVID-19 extracted from public sources. Contagion follows a compartmental epidemic model for each municipality; the latter, in turn, interact with each other through recurrent mobility. Considering the number of municipalities with confirmed COVID-19 cases, simulations can infer the level of mitigation (strong, moderate, or none) that each state is effectively adopting. Properties of the epidemic curves such as time and value of epidemic peak and outbreak duration have very broad distributions across different geographical locations. This outbreak variability is observed on several scales from state, passing through intermediate, immediate down to municipality levels. The epidemic waves start from several foci concentrated in highly populated regions and propagate towards the countryside. Correlations between delay of the epidemic outbreak and distance from the respective capital cities are strong in several states, showing propagation towards the countryside, and weak in others, signaling strong influences of multiple centers, not necessarily within the same state. Our take home message is that the responses of different regions to the same mitigation protocol can vary enormously such that the policies of combating COVID-19, such as quarantine or lockdown, must be engineered according to the region specificity but integrated with the overall situation. Even though we restricted our study to Brazil, we believe that these ideas can be generalized to other countries with continental scales and heterogeneous demographic distributions.

## I. INTRODUCTION

Since the coronavirus disease (COVID-19) outbreak was reported in Wuhan, Hubei province, China, at the end of December 2019, the scientific community has turned its attention to understanding and combating this novel threat [1–5]. A characteristic of this infectious disease, caused by the pathogen coronavirus SARS-CoV-2, is the high transmission rate of asymptomatic and preasymptomatic individuals [4, 6] who traveled unrestrictedly and spread COVID-19 through all continents, resulting in the pandemic declaration by the World Health Organization (WHO) on 11 March 2020. Efforts to track the transmission from Wuhan to the rest of mainland China and world has relied on mathematical modeling [2, 3, 5, 7] and was soon extended to other countries [8, 9], using the metapopulation approach [10–12]. In this modeling, the population is grouped in patches representing geographic regions and the epidemic contagion obeys standard compartmental models [13, 14] within the patches, while mobility among them promotes the epidemic to spread through the whole population.

The first case in Brazil was confirmed on 25 February 2020 in São Paulo, a case imported from Milan, Italy, the second epicenter of the epidemic, after Hubei province controlled COVID-19 with severe restrictions. On 5 March 2020 the first case was reported in Rio de Janeiro, next day in Feira de Santana and so on, spreading to all 27 federative units of Brazil. On 3 May 2020, the benchmarks of 100 000 confirmed cases with more than 7 000 deaths due to COVID-19 was surpassed in Brazil [15].

Brazil is a country of continental territory with an area of 8.5 × 10^6^ km^2^ demographically, economically, and developmentally very heterogeneous. Brazil is a federation divided into 26 states and one Federal District, its capital city. States are divided into municipalities; each has its own capital city. The total number of municipalities is 5570 [16]. According to estimates of 2019 [16], the total population of the country is 210 million. The most populated state is São Paulo with 46 million inhabitants and its homonymous capital city is the most populated municipality with 12.2 million inhabitants. The least populated state is Roraima, with 605 700. The least populated municipality is Serra da Saudade, in the state of Minas Gerais, with 781 inhabitants. Other characteristics of Brazilian demography are the broad distributions of the rural and urban population fractions and urban population densities of municipalities. Therefore, investigation of a pandemic and its consequences have necessarily to integrated the particularities of each region, which can be reckoned with a metapopulation approach.

In the present work, we investigate the epidemic of COVID-19 in Brazilian municipalities using a stochastic metapopulation model in which municipalities are represented by patches and contagious processes described by a SEAUCR compartmental model; see Fig. 1. Demographic and mobility data mined from public sources are used in the simulations. We intend to quantify how diversified the epidemic can be across the country rather than accurately predicting epidemic outbreaks in specific places. We use simulations to determine the level of dispersion in the epidemic curves in different places within a hypothetical scenario of uniform mitigation measures. We take the situation of 31 March 2020 as the initial condition, with foci in 410 municipalities, allowing to track the epidemics in a single integrated simulation in all municipalities. Our approach consists in calibrating the model with a few weeks of COVID-19 case data officially reported in Brazil and perform long-term analysis of epidemic outcomes using different mitigation scenarios. We find that the epidemic curves can vary enormously over scales raging from state to municipality levels. We also find that some states are characterized by outbreaks starting mainly in their capital cities, followed by epidemic waves propagating toward the countryside, while in other states there are multiple initial foci of epidemics. Our results provide basis for public policies to fight COVID-19 that should be simultaneously decentralized, in the sense that each locality has to consider its specificities, and integrated, since the fate of one region depends on the attitudes adopted by others.

**FIG. 1.**
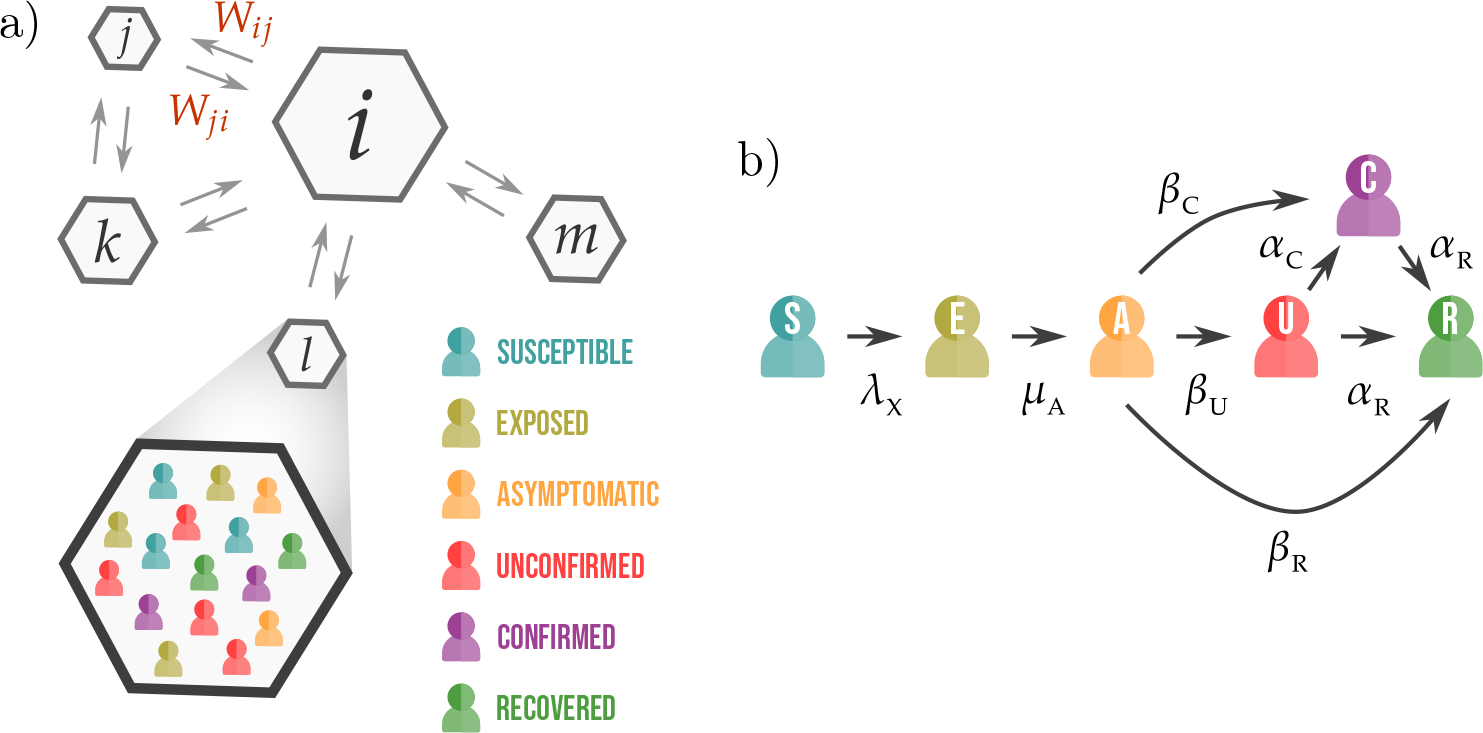
Schematic representation of the a) metapopulation structure and b) SEAUCR epidemic transitions with their respective rates. Patches *i* and *j* of different population sizes (represented by the areas) can interchange individuals with rates *W_ij_* and *W_ji_*. The epidemic events occur within patches under the hypothesis of well-mixed populations as illustrated in the zoom of patch *l*. Six compartments are used in the SEAUCR model: susceptible (S), exposed (E), asymptomatic (A), unconfirmed (U), confirmed (C) and recovered (R).

## II. THE MODEL

We study a stochastic metapopulation model for the spread of COVID-19 epidemics, in which a key ingredient is recurrent mobility [12] whereby individuals travel to other patches but return to their residence recurrently. The population is divided into patches which represent the places of residence as schematically shown in Fig. 1a). This model is inspired by a recent work [9] in which a microscopic Markov chain approach [17] was used to investigate the COVID-19 epidemic in Spain. A patch can represent a municipality, a neighborhood or any other demographic distribution of interest. Each patch is labeled by *i* = 1, 2,…, Ω, where Ω is the number of patches, and has a fixed resident population given by *N_i_*. The inter-patch (long distance) mobility rate is given by *W_ij_* defined as the number of individuals that travels from *i* to *j*, and return to *i* after a typical time *T*. Thus, 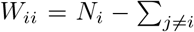 is the population of patch *i* that does not leave it. A patch *j* for which *W_ij_* > 0 is a *neighbor* of *i*. Different forms of mobility, with corresponding characteristic times, can be implemented. For the sake of simplicity, recurrent flux of individuals per day (*T*_r_ = 1 d) is used in the present work. Mobility within patches is not explicitly considered. Instead, we assume the well-mixed population hypothesis [13] where an individual has equal chance to be in contact with any other in the same patch.

The average number of contacts per individual of patch *i* per unit of time is *k_i_*. Variations in the social distancing can be represented by the *k_i_*. Contacts are related to local mobility and are strongly correlated with the population density ξ*_i_*. Monotonic increase has been proposed [18]. We follow Ref. [9] and use a contact number modulated by *k_i_* ∝ *f* (ξ*_i_*) with *f* (*x*) = 2 − exp(−*a*ξ). Due to the sparse demographic distribution of Brazil we used the urban population density of each municipality to compute ξ*_i_*. Interactions between urban and rural populations are also explicitly considered through a symmetric 2 × 2 contact matrix 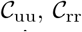, and 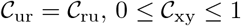, determining the contacts between urban (u) and rural (r) populations whose fractions are represented by *ω_i_* and 1 − *ω_i_*, respectively. The number of contacts in patch *i* is also modulated by

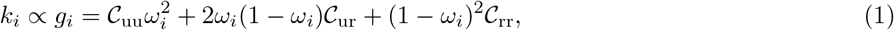

where the terms represent urban-urban, urban-rural, and rural-rural interactions from left to right. The contacts are parameterized by their average number per individual 〈*k*〉, such that

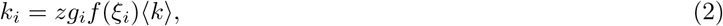

where *z* is a normalization factor, required by the relation 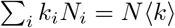 and given by

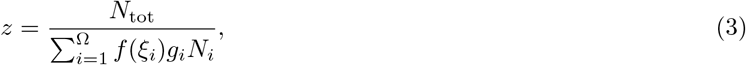

where 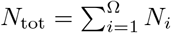 is the total population.

The epidemic dynamics within patches considers compartments susceptible (S), that can be infected, and recovered (R), i.e., immunized or deceased. Compartments of infected individuals are: exposed (E), that carries the virus but does not transmit it; asymptomatic (A), that have no symptoms or are presymptomatic but are contagious; unconfirmed (U), which are symptomatic but not yet identified by tests; and confirmed (C), that have tested positive for COVID-19. The epidemic transitions obey a SEAUCR model, in which susceptible individuals become exposed by contact with contagious persons (A, U, or C) with rates λ_A_, λ_U_, and λ_C_, respectively. Exposed individuals spontaneously become asymptomatic with rate *μ*_A_, while the latter transition to unconfirmed, confirmed, or recovered states with rates *β*_C_, *β*_U_, and, *β*_R_, respectively. Finally, unconfirmed or confirmed individuals are recovered (via recovery or death) with rate *α*_R_. The transition U→C has rate *α_C_* which depends on the number of unconfirmed cases 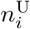. For a small 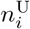 the confirmation rate is constant 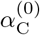, assuming that any municipality has the minimal resources for testing. The number of confirmed cases is limited by the finite testing capacity per inhabitant, ζ. We assume a simple monotonic function

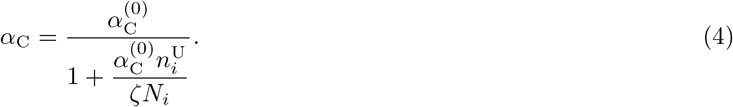

The transition E→U was not included since most studies agree that the asymptomatic or preasymptotic condition is a central characteristic of the COVID-19 epidemic [4, 9, 19]. The contact of confirmed individuals is reduced to *bk_i_* with *b* < 1. Figure 1b) shows a schematic representation of the epidemic events.

The population fraction moving recurrently from *i* to *j* is

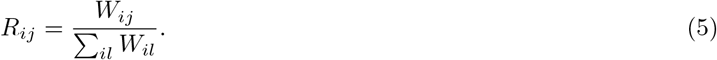

Thus, 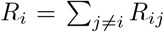 and *R_ii_* = 1 − *R_i_* are, respectively, the population fractions that move or not from patch *i*. Individuals residing in *i* can move to neighbors *j* following a stochastic process with rates *R_i_*/*T*_r_; the destination is chosen proportionally to *R_ij_*. Symptomatic (U or C) individuals do not leave their residence patches but if the transitions to U or C happen out of his/her residence they return with rate 1/*T*_r_, as do the individuals in the other states. Infection, healing and other epidemic state transitions follow Poisson processes with the corresponding rates. The details of the computer implementation, using Gillespie algorithm [20] are given in section IV of the Supporting Information (SI) [21]. Due to optimizations explained in the SI [21], simulations are viable for very large systems with reasonable computational resources. For example, an epidemic process lasting two years can be run in a few hours.

## III. THE PARAMETERS

### A. Mobility

Mobility parameters were extracted from public sources. Local commuting 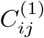 was estimated from the national census data of 2010 provided by the *Instituto Brasileiro de Geografia e Estatística* (IBGE) [22] as the number of people that daily commute among municipalities to work or study. We excluded from the dataset those travels corresponding to more than 250 km, which are reckoned by airline travel data. Commuting data lacking destinations were included, assuming proportionality to the complete-information portion of the dataset; see [23] for details. For the sake of generality, the short-range recurrent flux is a function 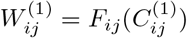. We use the simplest case *F_ij_*(*x*) = *x*. Air transportation data were obtained from the *Agência Nacional de Aviação Civil* (ANAC) [24] with the statistics of direct flights of 2014 considering the main nearby metropolitan region as the origin and destination of passengers (for example, Guarulhos, Viracopos and Congonhas airports were all linked to the municipality of São Paulo despite of being located in different municipalities). Using boarding and landing statistics of passengers, one-connection flights were inferred and the number of passengers 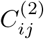 flying daily from *i* to *j* was estimated [25]. In order to construct a recurrent mobility with flights, we assume 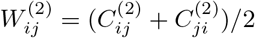, which represents an effective flux. However, the homogeneous mixing hypothesis within the patches means that only the number of individuals that stay part of the time in two different patches actually matters and not who traveled. The total recurrent mobility is 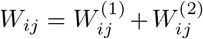; we assume that it occurs daily, i.e, *T*_r_ = 1 d.

### B. Epidemiological parameters

We used epidemiological parameters based on Ref. [9] which were mined from analyses of COVID-19 epidemic spreading on Hubei province China [1, 6, 8]. The time between the first contact with the pathogen and symptoms, that is the time in exposed and asymptomatic compartments, was taken as 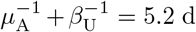 [1]. We used 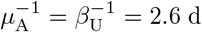. The recovering time of a symptomatic individual was estimated as 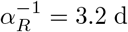 [6, 8]. For the asymptomatic individuals, we used the same recovering time of those who were symptomatic, such that 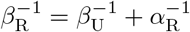. The infection rates λ_C_ = λ_U_ = λ_A_ = 0.06 d^−1^ [3, 26] and average number of contacts 〈*k*〉 = 13 were estimated to reproduce the scaling of Brazilian very initial exponential growth (~ exp0.3*t*) in reported cases of COVID-19; see Fig. SI-14a) of the SI [21]. These parameters provide an effective infection rate λ〈*k*〉 = 0.78 that represents an optimistic estimate nearer to the lower bounds estimated in other studies of COVID-19 spreading [6, 8, 9, 27]. The confirmation rate 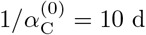 was calibrated together with the initial conditions (subsection IIID) to fit the amplitude after 31 March 2020 and also the total number of municipalities with confirmed cases in a moderate mitigation scenario; see subsection IV A. Up to 23 April 2020, Brazil had performed less than 1400 tests per million inhabitants [28]. Assuming a time window of 45 d for testing, we have approximately 1/30 000 per inhabitant a day. Since tests can be applied more than once in the same infected patient, we use ζ = 1/40 000 d^−1^ per inhabitant. This is a very rough estimate since the official number of tests is unknown at the moment of writing. Due to the testing targeted only to severe cases of respiratory syndromes adopted in Brazil to the date of investigation, the transition from asymptomatic to confirmed was neglected setting *β*_C_ = 0; we note that the model can easily include such a transition if the testing scenario changes.

### C. Contact parameters

Each patch represents a municipality with population given by the *Instituto Brasileiro de Geografía e Estatística* (IBGE) using estimates of 2019 [16], while the respective fractions of rural and urban populations were obtained from the IBGE census of 2010 [22]. The population density was determined using urban areas, obtained from high resolution satellite images, provided by *Empresa Brasileira de Pesquisa Agropecutíria* (EMBRAPA) [29]. These data are publicly available in the cited references. Both fraction and density of urban population have broad distributions as can be seen in Figs. SI-14b) and c) of the SI [21]. Therefore, the number of contacts in the municipalities estimated with Eq. (2) also has a broad distribution as shown in Fig. SI-14d) of the SI [21]. These are key features to enhance the accuracy of the prediction beyond large metropolitan centers. The contact of the individuals confirmed with COVID-19 was depleted to a fraction *b* = 0.3 of the regular contacts. Parameter *a* that controls the function *f*(*x*) was taken as *a* = 1/〈ξ〉 where 〈ξ〉 = 2791 inhabitants/km^2^ is the average urban population density of Brazil. The rural-urban contact matrix was chosen as 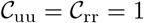 and 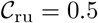. This last choice only assumes that urban-rural contacts are significantly smaller than urban-urban or rural-rural interactions.

### D. Initial conditions

The initial condition is a major hurdle for a detailed epidemic analysis due to high underreporting of cases for COVID-19. For the Wuhan outbreak it was estimated that undocumented infections were the source of at least 79% of documented cases [4], which include asymptomatic, preasymptomatic, and paucisymptomatic cases. This number can be larger depending on the testing policies [30, 31]. We adopt a simple linear correlation between number of confirmed cases reported by the official authorities and infected cases (E, A, and U). We use as the initial condition, 31 March 2020. Let 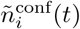 be the number of confirmed cases of COVID-19 in each municipality *i* [15] at day *t*, and let *t*_0_ correspond to 31 March and *t*_1_ to 4 April 2020. These data are provided in SI [21]. The initial condition at *t*_0_ is

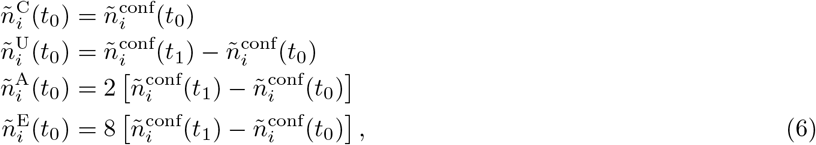

where 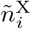 is the number of individuals in patch *i* and state X. This approach, based on the hypothesis that the increase of reported cases is strongly correlated with local transmission, is a way to reduce the effects of underreporting. The prefactors were chosen together with the parameter 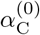 to fit both the initial increase (first two weeks of March) of the total number cases and of municipalities with confirmed cases under a moderate mitigation scenario; see subsection. IV A.

### E. Comments on parameter choices

The extensive set of parameters is aimed to reproduce the essence of the epidemic spreading at a country-wide level but not to track accurately the epidemic outbreak in every state or municipality. We specially assume uniform and constant control parameters for reduction of long-distance movement and contacts as well as the testing rates. This is surely not the case. Each federative state or municipality has its own mitigation and testing policies that are very heterogeneous. So, assuming uniformity and fitting the overall data at a country level, it is possible to infer in which regions the epidemic is evolving faster or slower than the average. Finally, our initial condition is a guess driven by data but still limited. The level of underreporting is known to be high, and broad ranges of values have been reported [4, 30, 31]. One limitation that can be easily identified is that municipalities without confirmed cases on 4 April are assumed to have no infected individuals of any type (E, A, U, or C) at March 31, which is probably not the case of the real system.

## IV. RESULTS

Different mitigation strategies are studied by uniformly reducing long distance mobility and contact by factors (*M, K*), i.e., *W_ij_* → (1 − *M*)*W_ij_*, for *j* ≠ *i*, and *k_i_* → (1 − *K*)*k_i_*. The strongest mitigation scenario that we investigated was (0.8,0.5), meaning the reduction of 50% in contacts and a small permeability between patches. We do not tackle the regime of suppression that can eradicate the epidemics because the current approach adopted in Brazil is mitigation. Averages were evaluated over 10 independent simulations.

### A. Nowcasting and model calibration

Figure 2a) compares the reported data and simulations for the number of confirmed cases in the first three weeks under different mitigation scenarios: none, moderate, and strong represented by (*M*, *K*) = (0, 0), (0.4, 0.3), and (0.8, 0.5) respectively. As explained in subsection III D, the moderate mitigation parameters, which are consistent with the estimates of the *Google Community Mobility Reports* for Brazil within this time window [32], reproduce the overall averages over the whole country. The metapopulation approach allows to investigate the epidemic at a municipality level [8, 9, 11]. Figure 2b) shows the number of municipalities with confirmed cases obtained in simulations with different mitigations and reports for Brazil between 1 and 22 April of 2020. Again, the moderate mitigation provides the best agreement with the reported data. Deviations start to appear after three weeks. No tuning with respect to specific places was used and, consequently, it does not reproduce accurately the number of reported cases for all federative states as expected, see SI-1 of the SI [21], due to the diversified testing and lockdown policies across different states in contrast with the uniformity hypothesis of the model. A long-term tracking of the reported cases requires a continuous update of new reported cases and parameters.

**FIG. 2.**
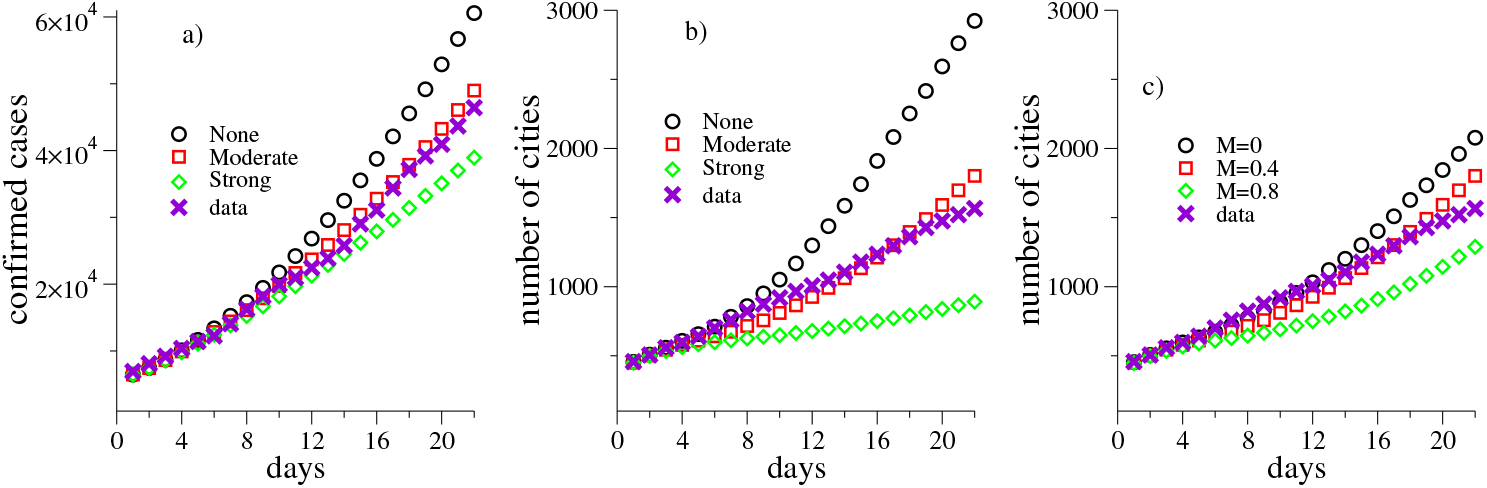
Comparison of simulations with reported data for COVID-19 in Brazil [15] for the first three weeks with day 0 corresponding to 31 March 2020. a) Number of confirmed cases and b) of municipalities with at least one confirmed case are presented. Different mitigation scenarios are considered in both a) and b): none with (*K,M*) = (0, 0), moderate with (*M,K*) = (0.3, 0.4), and strong with (*M,K*) = (0.2, 0.5). c) Evolution of number of municipalities with confirmed cases for a reduction of contact of *K* = 0.3 and different levels of long distance mobility *M* = 0, 0.4, and 0.8.

The role of long-distance traffic is investigated reducing the contacts by 30% and varying the mobility. The total number of cases changes very little since the leading contribution comes from local transmission in municipalities with larger incidence at the initial condition while the number of municipalities with confirmed cases, shown in Fig. 2c), depends significantly on the long-distance traffic restrictions. These results confirm the well know epidemiological finding that reducing long-distance transportation contributes to delaying the outbreak onset in different places, but alone alters the overall epidemic prevalence very little once the seeds have been spread [33], as already discussed for COVID-19 spreading on China [3, 34].

Irrespective of the heterogeneous testing approaches among states, we assume that essentially all municipalities have capacity to detect COVID-19 in the first hospitalized patients and, therefore, the number of municipalities with positive cases is expected to be an observable much less prone to the effects of underreporting. Figure 3 compares the evolution of municipalities with confirmed cases obtained in simulations with different mitigation measures and real data for each Brazilian state. The moderate mitigation parameters (*M*, *K*) = (0.4, 0.3), which fit better the overall data for Brazil, also perform well for most of the states in this time window and especially those with higher incidence of COVID-19, which lead the averages over the country. However, some states (AC, AM, PA, PE, PI, and RR - abbreviations for Brazilian federative states are given in Fig. 3) are more consistent with the simulations without mitigation while others (PB, PR, and RN) are nearer to a strong mitigation. A map with the classification of mitigation measures that best fit the data reported for each state is presented in Fig. 3.

**FIG. 3.**
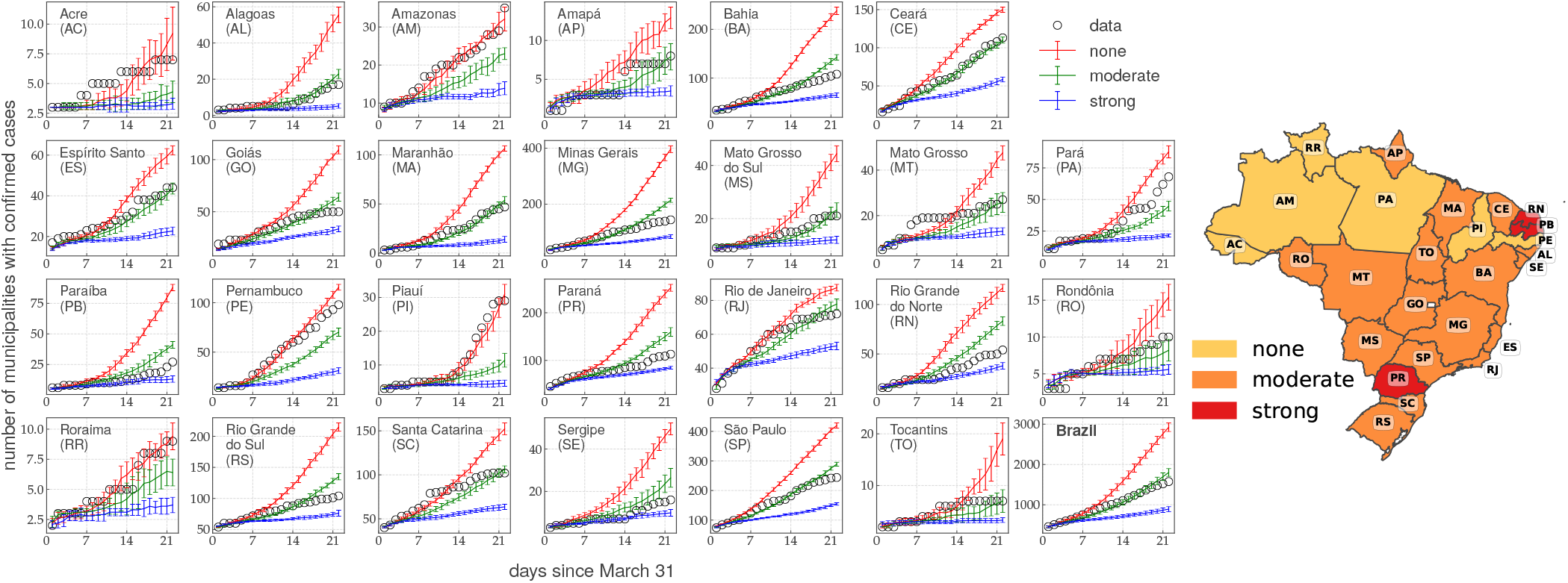
Simulations and observations of the number of municipalities with confirmed cases of COVID-19 for the 26 federative states of Brazil in the first three weeks of simulations. Data for the whole country are shown in the bottom right corner. Simulations were performed using none, moderate, and strong mitigations with parameters (*M*, *K*) = (0, 0), (0.4, 0.3), and (0.8, 0.5), respectively. Right: Map indicating the dominant mitigation scenario of each state for these 3 week simulations.

### B. Long-term analysis

We have shown that the model can be calibrated to work well for nowcasting, but forecasting is beyond the aim of this work due to large uncertainties of initial conditions, testing efficiency and dynamics of contact parameters. In forecasting, we should use nonuniform and varying parameters calibrated for each region. From now on, generic long-term predictions of the models are discussed while forecasting is left as future work.

Different mitigation scenarios are compared in Fig. 4a) where we present the fraction of symptomatic individuals, hereafter called epidemic prevalence, for the whole population. It confirms that suppressing long-distance mobility plays a minor role on the overall epidemic prevalence, only slightly reducing the maximum of the peak, as seen in the curves of *K* = 0.3 fixed and *M* varying. Cutting down the number of contacts is indeed the most efficient way to flatten the curve. The maximum number of symptomatic individuals drops by a factor 5 when the contacts are reduced to a half while the time of the epidemic peak doubles.

**FIG. 4.**
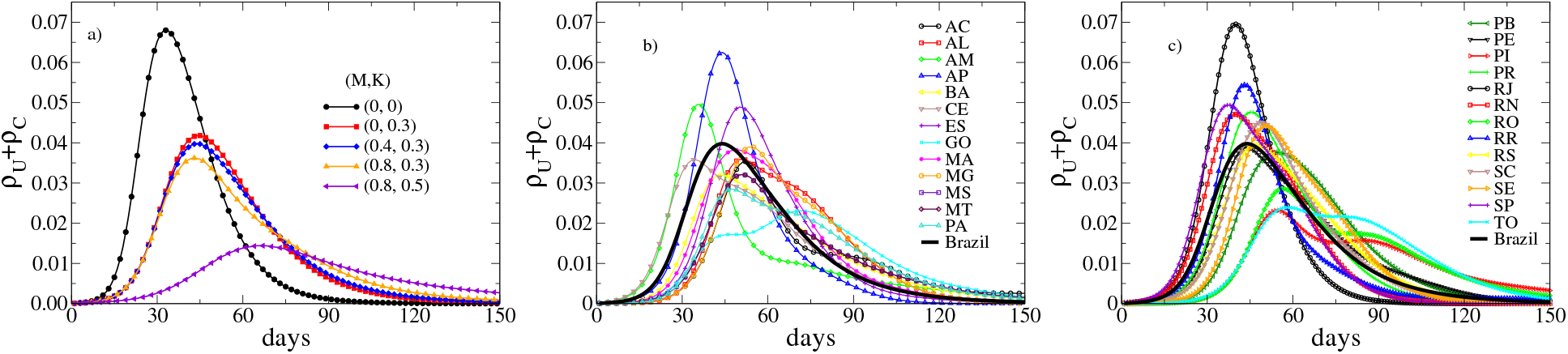
a) Evolution of the epidemic prevalence in the total population for different mitigation measures controlled by parameters (*M*, *K*). b,c) Epidemic prevalences for the 26 federative states of Brazil under a moderate mitigation approach with (*M,K*) = (0.4, 0.3). Day 0 corresponds to March 31, 2020.

Remarkable diversity of the epidemic outbreaks can be observed among all geographical scales. At the federative state level the time when the outbreak reaches the maximum epidemic prevalence, the epidemic peak, vary considerably as shown in Figs. 4b) and c), where the epidemic prevalence is shown as function of time for a moderate mitigation. Such a feature was observed in other metapopulation approaches for epidemic processes [8, 11]. Let *T* and *ρ* be the time and prevalence of the epidemic peak. According to the simulations with moderate mitigation, the first state to reach the peak would be CE (*T*_CE_ ≈ 34 d), followed closely by SP and AM (*T*_SP_ ≈ *T*_AM_ ≈ 36 d). The prevalences would be *ρ*_CE_ ≈ 3.6%, *ρ*_SP_ ≈ 4.9%, and *ρ*_AM_ ≈ 5.0%. The state with highest epidemic prevalence would be RJ with *ρ*_RJ_ ≈ 7.0% at day *T*_RJ_ ≈ 40. The epidemic peak happens later (*T*_MA_ = 71 d), with maximum less intense (*ρ*_MA_ ≈ 2.3%) and the curve broader for MA. The MA case is particularly interesting because while the peak for the state happens last, for its capital city São Luis it occurs at day 41 slightly earlier than average peak for the Brazil, which happens by day 44. In the simulations without mitigation, the times in days and prevalence percentage of the epidemic peaks are (*T*_CE_,*T*_SP_,*T*_AM_, *T*_RJ_,*T*_MA_) ≈ (28, 29, 26, 29, 48) and (*ρ*_CE_, *ρ*_SP_, *ρ*_AM_, *ρ*_RJ_, *ρ*_MA_) ≈ (5.9, 7.9, 7.5,10.9, 5.1) while in the case of strong mitigation simulations provide (*T*_CE_, *T*_SP_, *T*_AM_, *T*_RJ_, *T*_MA_) ≈ (49, 55, 56, 62, 68) and (*ρ*_CE_, *ρ*_SP_, *ρ*_AM_, *ρ*_RJ_, *ρ*_MA_) ≈ (1.5, 2.0, 2.2, 2.8, 0.6). Therefore, the model predicts that mitigation attitudes will lead to diverse levels of impact in different states even under the hypothesis of uniform measures in all regions. Table SI-II of the SI shows prevalence and time of the peaks for all states for these 3 mitigation strategies.

A more refined analysis can be done comparing immediate and intermediate geographical regions of a same federative state, which are determined by IBGE [35]. An immediate region is a structure of nearby urban areas with intense interchange for immediate needs such as purchasing of goods, search for work, health care, and education. An intermediate region agglomerates immediate regions preferentially with the inclusion of metropolitan areas [35] laying thus between immediate regions and federative states. In Brazil there are 133 intermediate and 510 immediate regions. Municipalities in different federative states do not belong to the same immediate regions even when they have strong ties. The broad variability of epidemic outbreaks observed at the federative state level is repeated at higher resolutions of geographical aggregation. Figure 5 shows the multi-scale structure of the outbreaks within MG, comparing the epidemic prevalence of different intermediate regions (top), followed by the next scale with all immediate regions belonging to the two intermediate ones presenting the epidemic peak first and last (middle), and finally the highest resolution with curves for municipalities (bottom) within the immediate regions with earliest and latest maxima shown in Figs. 5b) and c).

**FIG. 5.**
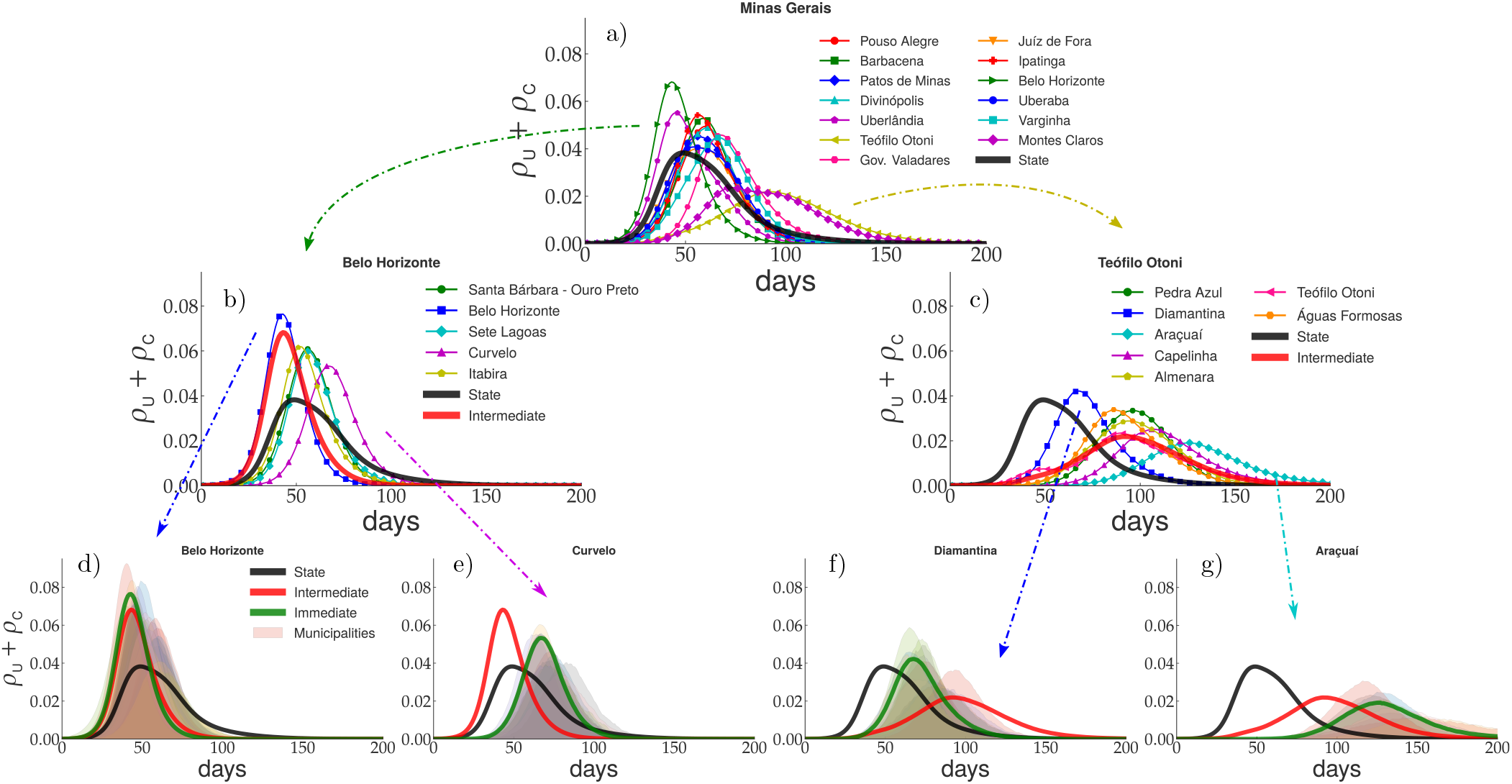
Multi-scale analysis of epidemic prevalence for MG state at several scales of geographical organization considering moderate mitigation with parameters (*M, K*) = (0.4, 0.3). Epidemic curves averaged with geographical resolution increasing from top to bottom are compared with lower resolution averages: a) intermediate regions, b,c) immediate regions, d-g) municipalities. The curves presenting the earliest and latest maxima are chosen as representative within each panel. Arrows indicate curves selected for zooming. Day 0 corresponds to 31 March 2020.

A large dispersion through different intermediate regions is seen in Fig. 5a), where the epidemic curves for the 13 intermediate regions of MG state are presented. The times and prevalence values at the peak differ by factors 2 and 6, respectively, between the regions that attain the maximum first and last, while the width of the curve is larger where the epidemics is more delayed. Curves for all intermediate regions of the 26 federative states are shown in Fig. SI-7 of the SI [21]. Stepping down to immediate regions, a similar behavior is found in Figs. 5b) and c). The intermediate and immediate regions of Belo Horizonte, that include the capital city and are by far the most populous regions of the state, lead the averages over state and intermediate regions, respectively, hiding the actual prevalence in other localities. Variability is still large within immediate regions as shown in Figs. 5d-g) with the epidemic prevalence (transparent curves) at different municipalities of the same immediate region. This pattern is repeated for other federative states: only the dispersion amplitude varies. Figures describing this multi-scale nature for CE, RJ, RS, and SP states are provided in Figs. SI-2, SI-3, SI-4, and SI-5, respectively, of the SI [21].

Variability of the time, prevalence, and width *τ*, which represents the outbreak duration, of the epidemic peak throughout immediate regions are shown in box plots of Fig. 6 for simulations with moderate mitigation. Box plots for none and strong mitigations are qualitatively similar and presented in Figs. SI-9 and SI-10 of the SI [21]. One can see that federative states with smaller territorial areas, such as SE, RN, and RJ, tend to have smaller dispersions of the peak time while those with larger scales and more sparsely connected territories, as AM, MT, and MS, have larger dispersions and outliers. We also have states with large but better connected territories such as SP, PR, and RS, for which the dispersion is significant but not characterized by outliers. Irrespective of the more cohesive pattern of outbreaks, the epidemic peaks in immediate regions of SP state can take more than twice as long as the peak at the capital city. On other hand, the outbreaks in RJ state are less desynchronized than in its neighbors, MG and SP, with the latest immediate region reaching the epidemic peak only 50% later than the capital, Rio de Janeiro.

**FIG. 6.**
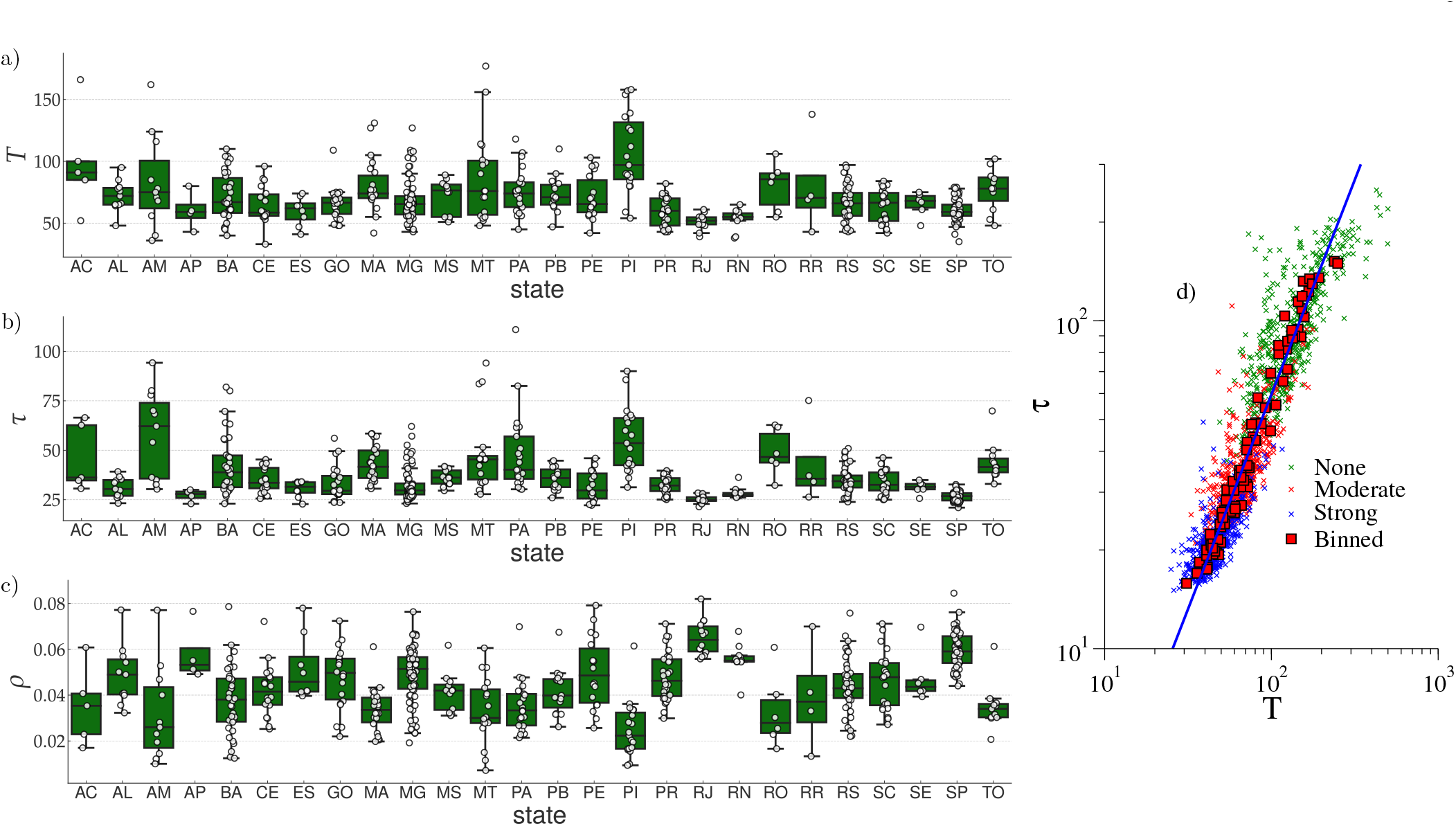
Box plot for the peak a) time *T* b) width *τ*, and c) prevalence *ρ* averaged over immediate regions for the 26 federative states using a moderate mitigation with parameters (*M,K*) = (0.4, 0.3). Circles are data for different regions. As usual, boxes yield median, lower and upper quartiles while points outside whiskers are outliers. d) Scatter plot for peak width versus time in days since 31 March2020 for three mitigation scenarios: none, moderate and strong. Squares are the averages binned over federative states for each mitigation method while the line is a power-law regression of the binned data.

Another interesting observation in Fig. 6 is that the more delayed the epidemic peak, the lower the maximum prevalence and the longer its duration. For the susceptible-infected-recovered (SIR), the simplest compartmental model with immunity, these relations can be analytically derived if the prevalence is not too large and asymptotically provides *ρ* ~ *T*^−2^ and *τ* ~ *T* [14] (~ means asymptotic proportionality). A scatter plot of peak width *τ* versus time *T* for distinct immediate regions and mitigation approaches is shown in Fig. 6d). Power-law relations with exponents *τ* ~ *T*^1.3(1)^, Fig. 6d), and *ρ* ~ T^−1.9(2)^ (not shown) are obtained. While the relation between peak value and time is consistent with simple compartmental models, the outbreak duration increases superlinearly with *T*. The outbreak size can be estimated as *N*_out_ ≃ *ρ*τ ~ *T*^−1^ for standard SIR while *N*_out_ ~ *T*^−0.6^ in our metapopulation simulations, indicating that the reduction of total number of infected individuals on places with delayed outbreaks will be proportionally lower than the prediction of simple compartmental models. This can be rationalized in terms of multiple seeding of epidemics due to flux of people from other municipalities.

The space-time progression of the epidemics presents complex patterns as shown in Figure 7, where color maps with the prevalences of symptomatic cases in each municipality in Brazil are shown within at three-weeks intervals for moderate mitigation simulations. Full evolutions for this and other investigated parameters are provided in Videos SI-1, SI-2, SI-3, and Figs. SI-12 and SI-13 of the SI [21]. An average epidemic wave propagating approximately from the east, where the most populated regions are found, to northwest can be seen at a country level. Moreover, when the epidemic is losing strength in the east, the countryside is still facing high epidemic levels. Maps and movies suggest that the epidemic starts in the more populated places, usually the largest metropolitan regions, and spreads out towards the countryside. This is quite clear in SP, where the focus begins in its capital city, and moves westward into the countryside. However, other states as PR and RS follow a different pattern with multiple foci evolving simultaneously.

**FIG. 7.**
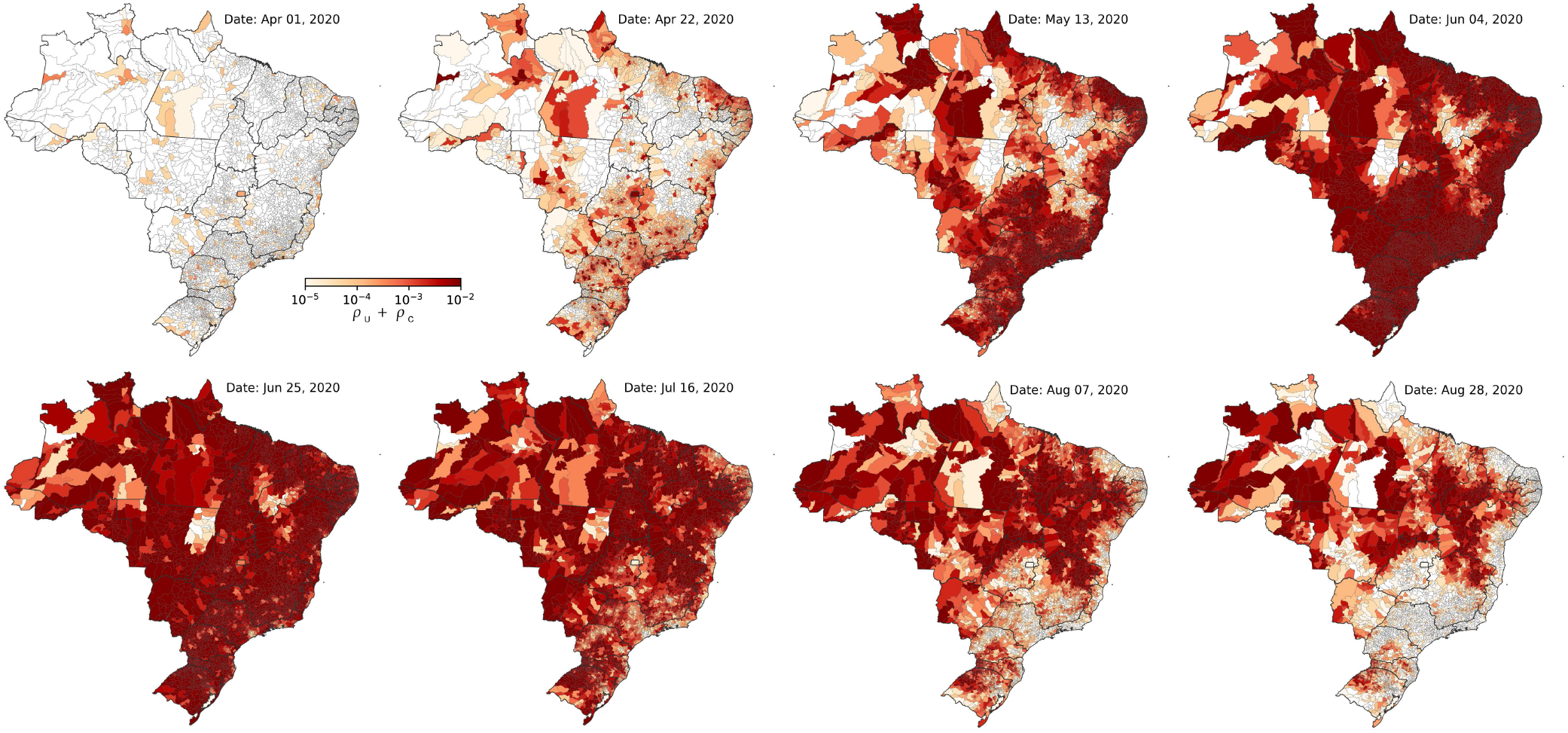
Color maps presenting the evolution of the prevalence of symptomatic cases (U and C) for Brazil in a simulation with a moderate mitigation using parameters (*M,K*) = (0.4, 0.3).

We analyzed the correlation between time *T* of the epidemic peak in immediate regions and their distances *D* from the capital city of the respective state. Figures 8a)-c) show scatter plots of *D* vs *T* for three typical correlation patterns found in simulations. The remaining states are given in Fig. SI-11 of the SI [21]. SP presents a strong correlation with Pearson coefficient *r* = 0.69, statistically significant with *p*-value less than 10^−7^. For PR, no statistical correlation is observed. Finally, MG has a moderate correlation with Pearson coefficient *r* = 0.47, but statistically significant with *p*-value of 10^−5^. We classified the states according to the correlation between *T* and *D* in three groups: *not significant* for which the *p*-value is larger than 0.02; *strong correlation* that has statistical significance with *p* < 0.02 and *r* > 0.6; and *moderate correlation* with *p* < 0.02 and *r* < 0.6. The results are summarized in Fig. 8d) where the map shows states colored according to their classification. The structure does not change for other mitigation approaches. The complete set of Pearson coefficients and *p*-values are given in Table SI-I of the SI [21]. States in the north (RR, RO, AP, AC, AM, PA, and TO) have few immediate regions distributed in large territories and are less connected. Thus, lack of correlations is not surprising. Moderate correlations in such states as MG, BA, and GO, with large but better connected territories are due to the existence of multiple important regions that play the role of regional capitals. However, PR is an extreme case with many and better connected immediate regions but total lack of correlation with the capital city, Curitiba. Indeed, PR has economically and culturally independent regions such as Londrina and Maringa with strong ties with SP, and the touristic region of Foz do Iguacu with Paraguay and Argentina.

**FIG. 8.**
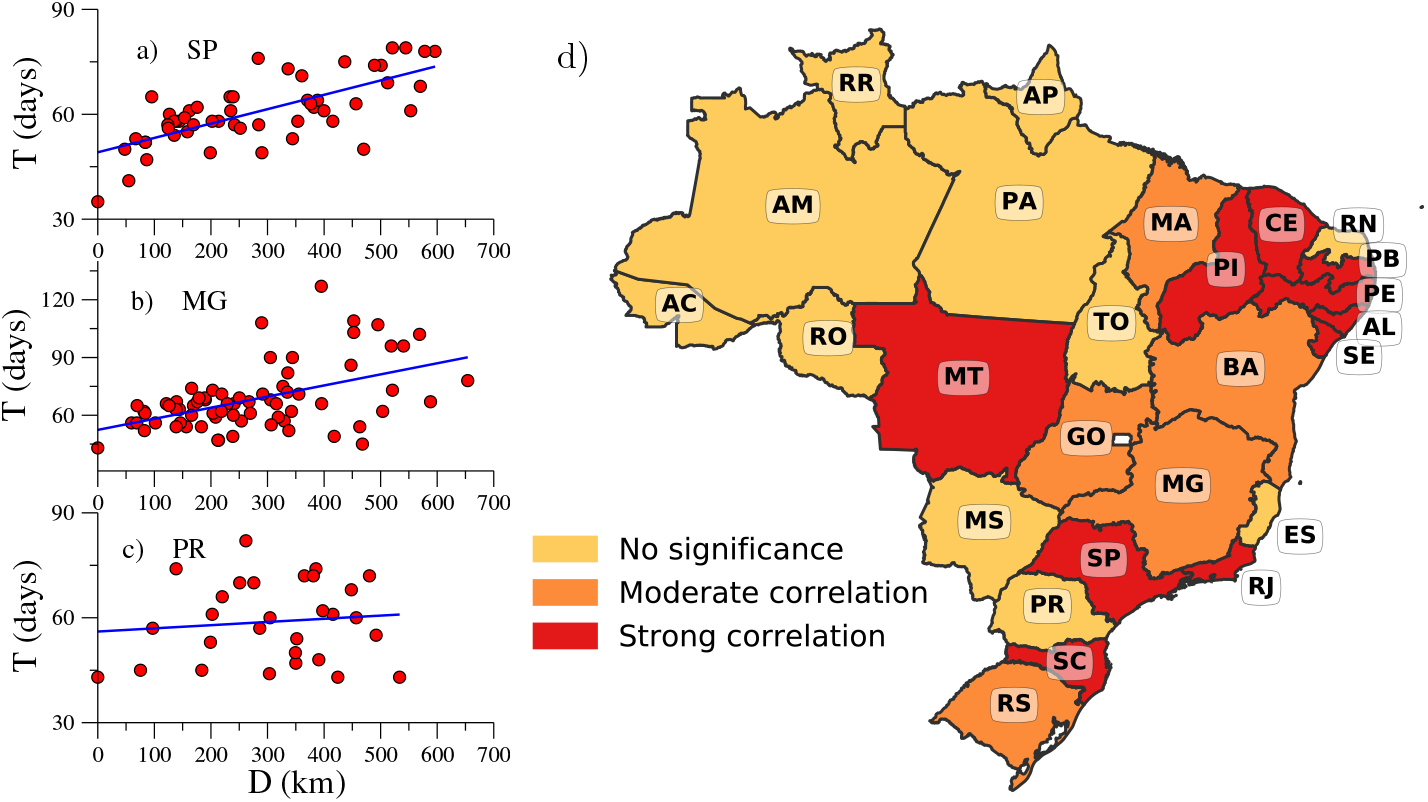
Scatter plots with time for the peak *T* and distances of immediate regions to capitals *D* of the respective a) SP, b) MG and, c) PR states. d) Map with the classification of typical correlation between *T* and *D* of each federative state. See main text for criteria.

## V. DISCUSSION

The wide territorial, demographic, infrastructural diversity of large countries such as Brazil demands modeling of the COVID-19 epidemic with geographical resolution higher than usual compartmental epidemic models, which can be achieved using the metapopulation framework. Performing simulations of at the municipality level for Brazil with diverse mitigation scenarios (none, moderate, and strong) and considering the integration of different regions through long-distance mobility of persons, we have identified a high degree of heterogeneity and desynchronization of the epidemic curves in the main metropolitan areas and countryside regions. The diversity of outcomes is observed in several geographical scales from states to immediate regions, that encloses groups of nearby municipalities with strong economic and social ties. The stronger the mitigation attitudes, the more remarkable are the differences. We find moderate or strong correlation between the delay of the epidemic peak in countryside regions when compared with the capital cities of the respective state for most states with well connected immediate regions. However, one exception is PR where diverse epidemic foci evolving apparently independently of the capital city.

The administrative organization of Brazil gives to the federative states and municipalities some independence to adopt mitigation approaches, but these are limited by economic dependence on superior spheres. So, the tendency is to have similar approaches within states. Our simulations indicate that uniform mitigation measures may not be the optimal strategy. In a municipality where the peak naturally happens after the capital of its state, it will delay even more if strong mitigation measures are adopted synchronously with the capital. On the one hand, such a municipality probably would have to extend the mitigation for much longer periods since once the epicenters of epidemics start to relax their restrictions, the viruses will circulate faster, reaching these vulnerable municipalities with essentially the entire population still susceptible. The social and economic impacts could be higher. On the other hand, an eventual collapse of the local heath system in countryside regions could also occur after the larger metropolitan areas are under control, raising the possibility of unburdening the local healthy care system.

In fact, the desynchronization of the epidemic curves and consequently of the health-system collapse may be an asset in designing optimal resource allocation. One important attitude is to preserve these reservoirs in countryside as long as possible. This could be done in at least two ways. The first is reducing to the lowest possible level the flux of people from areas with high incidence of COVID-19, in other words, strict sanitary cordons in the epidemic epicenters. This approach is theoretically obvious but can hardly be achieved in practice and only Wuhan had success in this endeavor [5]. The second option is to flatten the curves as much as possible in the main metropolitan regions. Beyond the positive consequences for their own health systems, this attitude would have the positive side effect of delaying outbreaks in the countryside. Our study shows that countryside regions are not safe in the medium-term but have precious additional time to prepare. Not less important, constant monitoring of possible local transmission by massive testing policies [36] even without significant epidemic incidence, is still the only safe path to keep the countryside economically active while other epidemic epicenters are locked down.

## Data Availability

Data will be available after peer-review.

https://covidbr.github.io/

## ACKNOWLEDGMENTS

We thank Marcelo F. C. Gomes for providing the filtered mobility data used in the present work; Ronald Dickman for useful discussion and manuscript reading; and Jesus Góméz-Gardeñes and Alex Arenas for discussions that inspired the beginning of this work. WC thanks David Soriano-Paños for extensive discussions about metapopulation modeling and Mario Balan for discussions about IBGE data. WC thanks the kind hospitality of University of Zaragoza and GOTHAM Lab during the realization of the work. SCF is in profound debit with Zélia Almeida who kept two children calm and safe under the same roof where this work was performed during the quarantine. This work was partially supported by the Brazilian agencies CNPq and FAPEMIG. This study was financed in part by the Coordenação de Aperfeicoamento de Pessoal de Nivel Superior - Brasil (CAPES) - Finance Code 001.

## Author contributions

GSC performed the stochastic simulations. WC and GSC prepared the figures and movies. SCF wrote the first version of the manuscript. All authors contributed equally to result analysis, manuscript revision, and research conception.

## Competing interests

All authors declare no competing interest.

## Data availability

All data used in our simulations are available in public repositories [15, 22, 24, 29].

## Corresponding author

Silvio C. Ferreira.

## Notes

### Competing Interest Statement

The authors have declared no competing interest.

